# Overall burden and characteristics of COVID-19 in the United States during 2020

**DOI:** 10.1101/2021.02.15.21251777

**Authors:** Sen Pei, Teresa K. Yamana, Sasikiran Kandula, Marta Galanti, Jeffrey Shaman

**Affiliations:** Department of Environmental Health Sciences, Mailman School of Public Health, Columbia University, New York, NY 10032, USA

## Abstract

The COVID-19 pandemic disrupted health systems and economies throughout the world during 2020 and was particularly devastating for the United States. Many of epidemiological features that produced observed rates of morbidity and mortality have not been thoroughly assessed. Here we use a data-driven model-inference approach to simulate the pandemic at county-scale in the United States during 2020 and estimate critical, time-varying epidemiological properties underpinning the dynamics of the virus. The pandemic in the US during 2020 was characterized by an overall ascertainment rate of 21.6% (95% credible interval (CI):18.9 – 25.5%). Population susceptibility at year’s end was 68.8% (63.4 – 75.3%), indicating roughly one third of the US population had been infected. Community infectious rates, the percentage of people harboring a contagious infection, rose above 0.8% (0.6 – 1.0%) before the end of the year, and were as high as 2.4% in some major metropolitan areas. In contrast, the infection fatality rate fell to 0.3% by year’s end; however, community control of transmission, estimated from trends of the time-varying reproduction number, *Rt*, slackened during successive pandemic waves. In the coming months, as vaccines are distributed and administered and new more transmissible virus variants emerge and spread, greater use of non-pharmaceutical interventions will be needed.

---

The novel respiratory virus, severe acute respiratory syndrome coronavirus 2 (SARS-CoV-2), the causative agent of coronavirus disease 2019 (COVID-19) emerged in late 2019 in China^1,2^. The virus quickly spread around the world due to a combination of epidemiological features that enabled efficient transmission and geographic translocation: limited pre-existing sterilizing immunity in the human population^3^, sustained human-to-human transmission due to a reproduction number well above unity^4^, presymptomatic and asymptomatic contagiousness^5-7^, and a high proportion of undocumented infections^5^. By the end of 2020, more than 82.3 million cases and 1.8 million deaths had been reported worldwide^8^. While several vaccines conferring protection against the virus were approved and began to be distributed by the end of 2020^9,10^, the pandemic continues at record levels during early months of 2021.

During 2020, the United States (US) documented more COVID-19 cases and deaths than any other country in the world^8^. The first COVID-19 case was identified in Washington state on January 20, 2020. Over the course of the year, three pandemic waves took place: 1) a spring season outbreak in select, mostly urban areas following the introduction of the virus to the US; 2) a summer wave that predominantly affected the southern half of the country; and 3) a fall-winter wave that remains pervasive. To understand the transmission of the virus and better control its progression in the future, it is vital that the epidemiological features that have supported these outbreaks be quantified and analyzed in both space and time.

Here we use a county-resolved metapopulation model to simulate the transmission of SARS-CoV-2 within and between the 3142 counties of the US. The model depicts both documented and undocumented infections and is coupled with an iterative Bayesian inference algorithm—the ensemble adjustment Kalman filter—which assimilates observations of daily cases in each county and population movement between counties^11,12^ (see Supplementary Information). The Bayesian inference supports a fitting of the model to case observations and estimation of unobserved state variables (e.g. population susceptibility within a county) and system parameters (e.g. the ascertainment rate in each county). The model fitting captures the 3 waves of the outbreak as manifest at national scales (Fig 1a), as well as in major metropolitan areas throughout the country (Fig S1).

**Figure 1.**
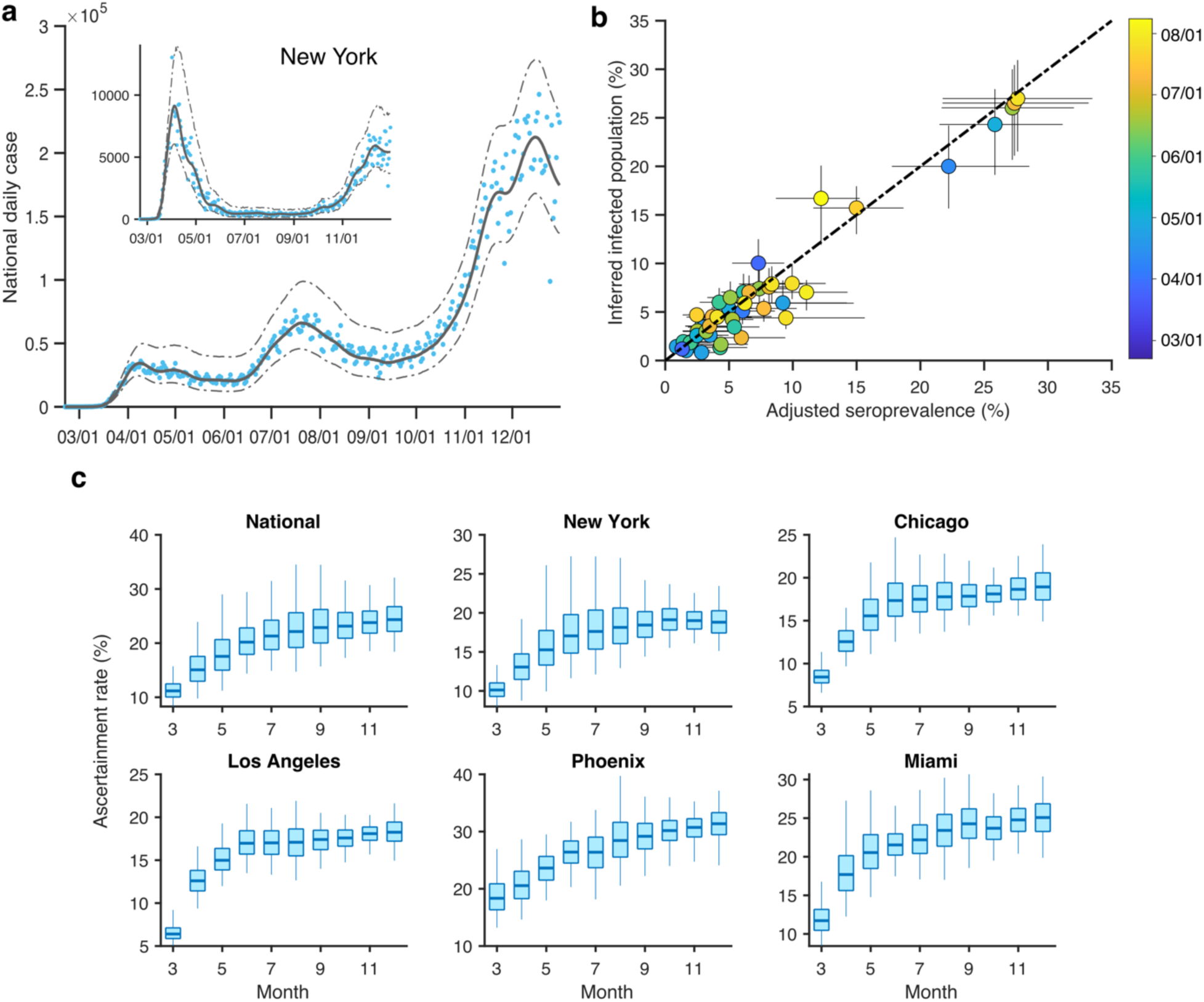
Model calibration and ascertainment rate. (a), Model fitting to daily case numbers (blue dots) in the US and the New York metropolitan area (inset). Solid and dashed lines show the median estimate and 95% CIs, respectively. (b), Comparison between inferred percentage cumulative infections and seroprevalence in 10 locations adjusted for antibody waning. Whiskers show 95% CIs, and color indicates the sample collection date in each location. Details on the serological survey are provided in Supplementary Information. (c), Distributions of estimated ascertainment rate in the US and five metropolitan areas. Boxes show median and interquartile range, and whiskers show 95% CIs. Monthly posterior estimates are presented for March to December 2020.

To further validate the simulations, we compared model estimates of cumulative infections to findings from US Centers for Disease Control and Prevention (CDC) seroprevalence surveys conducted at site and state levels^13^. The seroprevalence data, which provide an out-of-sample corroboration of the model fitting, were adjusted for the waning of antibody levels following adaptive immune response^14,15^ (see Supplementary Information, Figs S2-S3). Model estimates of cumulative infected percentages are well aligned with adjusted seroprevalence estimates from the CDC 10-site survey across sites and through time (Pearson r=0.97, mean absolute error (MAE)=1.34%) (Fig 1b) and are similarly well matched to adjusted estimates at the state level (Fig S4).

A critical feature of SARS-CoV-2 is its ability to infect and transmit largely from individuals never diagnosed with the virus^5^. The model structure and fitting enable estimation of the ascertainment rate, the percentage of infections confirmed diagnostically, at county scales. The national population-weighted ascertainment rate averaged for all of 2020 was 21.6% (95% credible interval (CI): 18.9 – 25.5%). This national ascertainment rate increased from 11.2% (8.3 – 15.7%) during March 2020 to 24.3 % (18.4 – 32.1%) during December 2020 (Fig 1c). The increase through time is a likely by-product of increasing testing capacity, a relaxation of initial restrictions on test usage, and increasing recognition, concern and care-seeking among the public. We additionally focus on 5 metropolitan areas within the US. Small differences in the ascertainment rate manifest across these areas; in particular, ascertainment rates for Phoenix and Miami are higher than the national average for much of the year, whereas New York City, Chicago, and Los Angeles are consistently below the national average.

At the national level, three pandemic waves are clearly evident during spring, summer and fall/winter (Fig 1a); however, among the 5 focus metropolitan areas the structure differs with New York and Chicago experiencing strong spring and fall/winter waves but little activity during summer, Los Angeles and Phoenix undergoing summer and fall/winter waves, and Miami experiencing all 3 waves (Fig S1). Los Angeles county, the largest county in the US with a population of more than 10 million people, was particularly hard hit during the fall/winter. The differences in virus activity produced different cumulative infection numbers through time (Fig 2a). Population susceptibility at the end of the year was 68.8% (63.4 – 75.3%) for the US, and among the focal metropolitan areas ranged from 47.9% (39.6 – 54.9%) in Los Angeles to 71.4% (65.4 – 74.2%) in Phoenix. Though there is variability among counties, a substantial portion of the US population (68.8%) had not been infected by the end of 2020; however, pockets of lower population susceptibility, which are evident in the southwest and southeast on August 1st (Fig. 2b), expanded considerably by December 1st (Fig 2c). In particular, areas of the upper Midwest and Mississippi valley, including the Dakotas, Minnesota, Wisconsin and Iowa, are estimated to have population susceptibility below 40% as of December 1, 2020.

**Figure 2.**
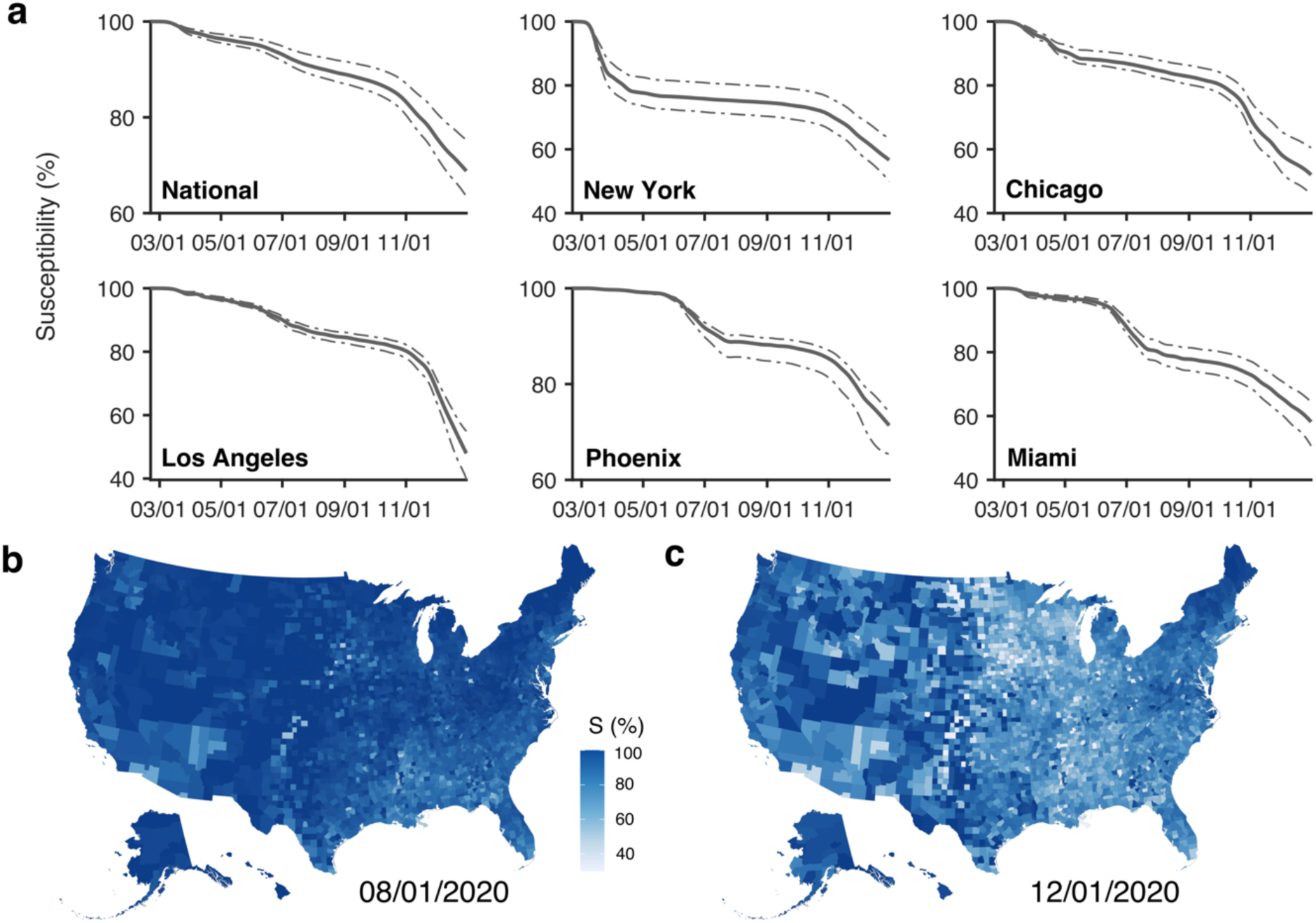
Estimates of population susceptibility. (a), Estimated evolution of susceptibility to COVID-19 in the US and five metropolitan areas. Solid and dash lines show median estimate and 95% CIs. (b-c), Estimated susceptibility in 3,142 US counties on August 1^st^ and December 1^st^ 2020. Color shows median estimate.

The structure of the outbreak is evident in both incidence and prevalence estimates (Figs 3, S5, S6). Incidence indicates the daily number of newly infectious individuals—both those who will be confirmed COVID-19 cases and those whose infections will remain undocumented. The majority of infections each month are undocumented (Fig 3a), as indicated by the low ascertainment rates (Fig 1c). For all of 2020, an estimated 78.4% of infections in the US were undocumented. Estimates of daily prevalence provide a measure of the community infectious rate, the fraction of the population currently harboring a contagious infection. National SARS-CoV-2 prevalence increased to 0.78% (0.61 – 1.00%) by December 31, indicating that roughly 1 in 128 persons was contagious (a similar percentage, 0.84% (0.53 – 1.28%), was estimated to be latently infected, i.e. infected but not yet contagious) (Fig 3b). Among the 5 focal metropolitan areas, prevalence varied considerably: in mid-November, Chicago reached a prevalence of 1.50% (1.28 – 1.77%); whereas prevalence in Miami rose to 1.22% (1.00 – 1.46%) during July. Los Angeles was even more burdened at the end of 2020 with a prevalence of 2.40% (2.00 – 2.89%) as of 31 December (Fig S6).

**Figure 3.**
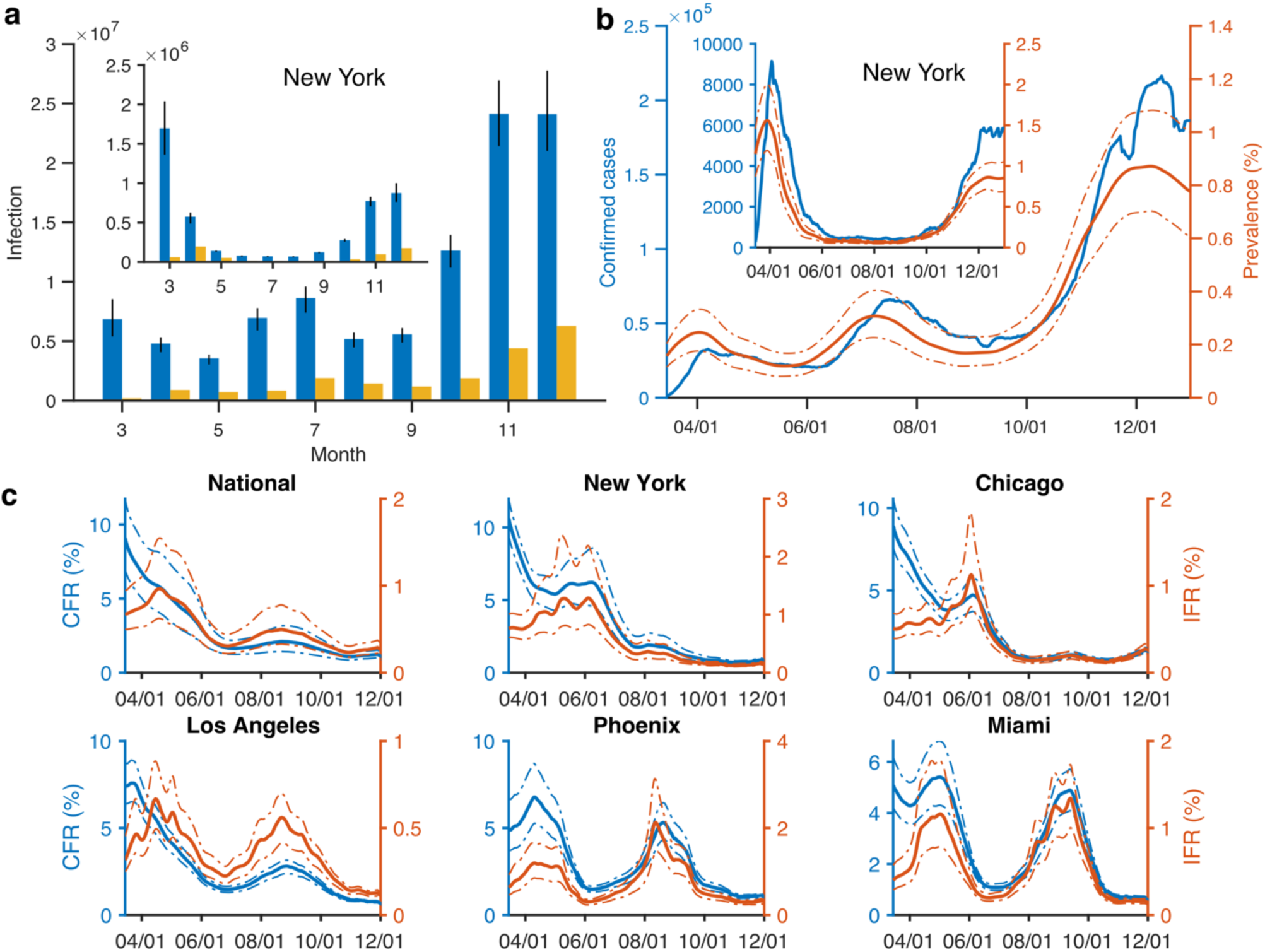
Estimated transmission and characteristics of COVID-19 in the US. (a), Estimated monthly total infections (blue bars, whiskers show 95% CIs) and confirmed cases (orange bars) in the US and the New York metropolitan area (inset). (b), Daily confirmed cases (blue line, 7-day moving average) and estimated prevalence of contagious infections (red line, median and 95% CIs) in the US. Result for the New York metropolitan area is shown in the inset. (c), Estimated CFR (blue lines) and IFR (red lines) in the US and five metropolitan areas. Solid and dash lines show median estimate and 95% CIs.

The model fitting enables estimation of the case fatality rate (CFR) and the infection fatality rate (IFR). Both rates were highest at the national level at the beginning of the spring wave: the CFR was 8.83% (6.75 – 11.40%) and the IFR was up to 0.96% (0.62 – 1.53%) in March and April (Fig 3c). Over the course of the year, with earlier diagnosis and treatment, improved patient care, and, in the case of CFR, increased reporting of mild infections, the CFR and IFR dropped to 1.16% (0.95– 1.48%) and 0.28% (0.22 – 0.35%) by December, respectively. Both rates varied by location and over time; for instance, intermediate drops of CFR and IFR began for Los Angeles, Phoenix and Miami prior to the summer wave in association with a decrease in the average age of documented infection (Fig S7). Overall, these findings delineate the mortality risk associated with infection broadly. The national IFR during the latter half of 2020 hovers around 0.30%, well above estimates for both seasonal influenza (<0.02%)^16^ and the 2009 influenza pandemic (0.0076%)^17^.

Given the high numbers of cases and deaths, and the successive pandemic waves, a central question is whether local populations within the US have responded to the growth of infections in their communities with improved control through non-pharmaceutical interventions (NPIs). Such control, effected through mask usage, social distancing, indoor ventilation, surface cleaning and restrictions on mass gatherings and other indoor activities, is reflected in the modulation of the time-varying reproduction number, *R*_*t*_. A decrease of *R*_*t*_ over time suggests a community is improving control of the virus by regulation or public adoption of control measures. For each of the three waves during 2020, we identified counties that experienced 2 or more consecutive weeks of increasing (or decreasing) reported cases and also reported 15 daily cases per 100,000 persons at least once during the period. We then examined trends of *R*_*t*_ for each of these 3 periods. During the earliest period in the spring, *R*_*t*_ decreased when counties experienced case growth and case decline; however, the decline in *R*_*t*_ is more precipitous when counties experienced growth (Fig 4a). During the second, summer period, the decline of *R*_*t*_ in counties with case growth starts at a lower level and is more muted, and counties experiencing declining case numbers instead have an increase of *R*_*t*_.

**Figure 4.**
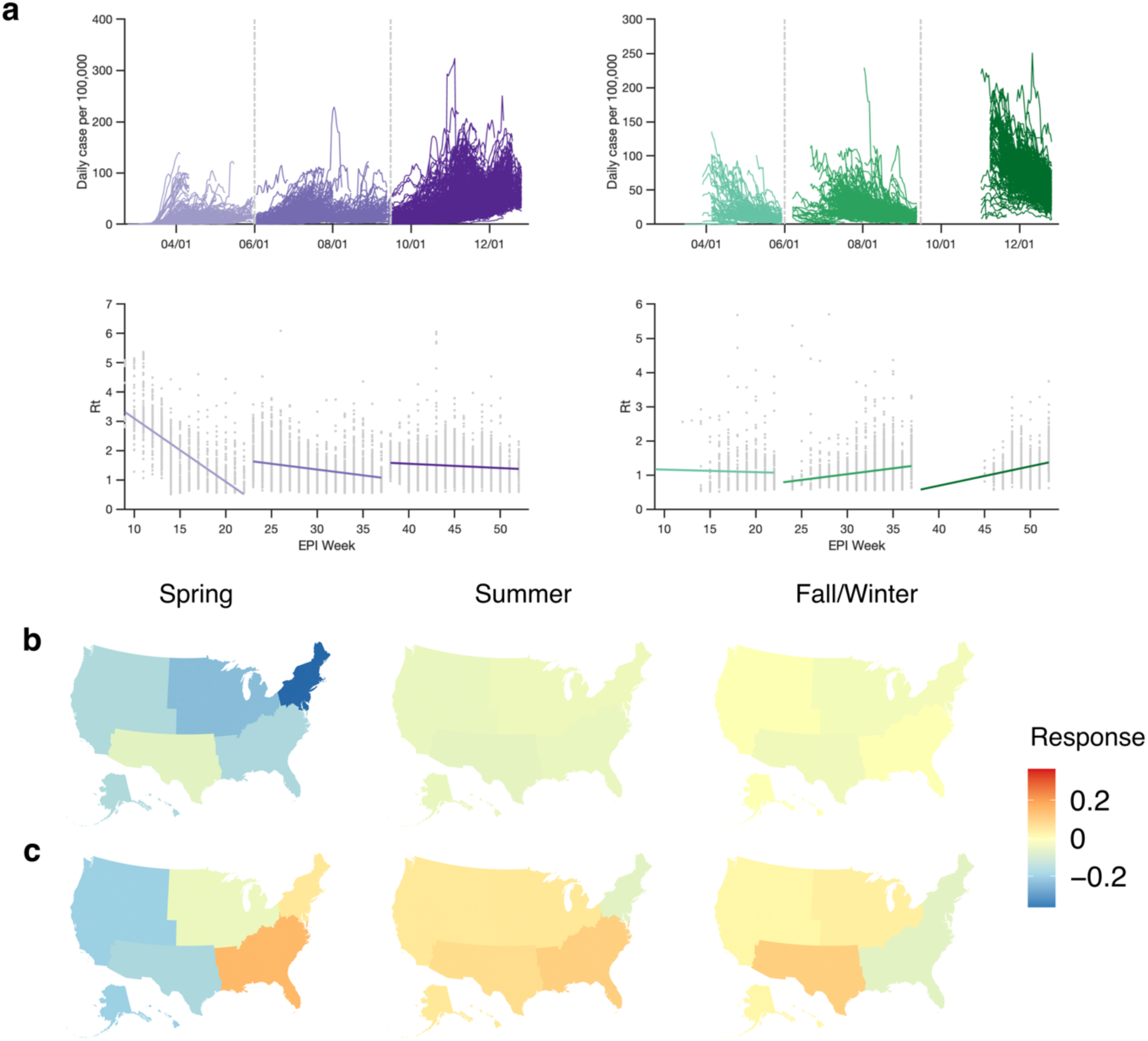
Response to growth and decline of local infections. (a), County-level curves of daily confirmed cases (per 100,000 people) that increase (top left) or decrease (top right) for at least two weeks. Vertical dash lines separate the spring, summer and fall/winter waves. Weekly reproduction numbers R_t_ in counties with increasing (bottom left) or decreasing (bottom right) local infections are reported. Solid lines are linear fit. The x-axis shows the epidemiological weeks used by the US CDC (EPI week). (b-c), Response (slope of linear fit) to increasing (top) and decreasing (bottom) local infections during the spring (left), summer (middle) and fall/winter (right) waves. Results are shown for five US regions: Northeast, Southeast, Midwest, Southwest, and West.

During the final fall/winter period, the patterns are similar to the summer period with *R*_*t*_ decreasing in counties experiencing case growth but decreasing in counties experiencing case decline. The same trends are present when averaging the counties to regional scales (Fig 4b,c), as well as when using different case per capita thresholds (Figs. S8-S9). Overall, this analysis suggests a lessening ability or willingness to control SARS-CoV-2 in the US as the year progressed. Part of this year-long trend, particularly during the fall/winter wave, is likely due to seasonal effects that moderate *R*_*t*_ and are beyond direct human control. In particular, evidence suggests that SARS-CoV-2 is more transmissible when humidity levels are low^18^, as is the case during winter in temperate regions, and that people are bound to spend more time indoors during winter when temperatures are low. Both effects likely increased opportunities for transmission during the third wave and for the most part cannot be effectively counteracted. However, policies and behaviors, specifically the use of masks, social distancing, ventilation, restricting mass gatherings and indoor dining, etc., that limit opportunities for virus transmission are subject to regulation and individual choice. Individual control behaviors may have slackened towards the end of the year, and policies allowing indoor dining and other commercial activities were more common late in the year^19^. The relative contributions of seasonal versus behavioral effects on changing *R*_*t*_ trends across successive pandemic waves cannot be disentangled in this analysis.

The US experienced the highest numbers of COVID-19 cases and deaths in the world during 2020. Our findings provide quantification of the time-evolving epidemiological characteristics associated with successive pandemic waves in the US, as well as conditions at the end of the year and prospects for 2021. Critically, despite more than 19.6 million reported cases at year’s end, more than 68% of the population remained susceptible to viral infection. Several factors will considerably alter population susceptibility in the coming months. Firstly, ongoing transmission will infect naïve hosts and continue to deplete the susceptible pool. Secondly, as more vaccine is distributed and administered, more individuals will be protected against symptomatic infection and the IFR will decrease. Lastly, our model does not represent re-infection, either through waning immunity or immune escape; however, re-infection has been documented^20,21^, evidence of waning antibody levels exists^22,23^, and new variants of concern have emerged^24,25^ and will likely continue to do so. All these processes will affect population susceptibility over time and help determine when society enters a post-pandemic phase, the pattern of endemicity the virus ultimately assumes, and its long-term public health burden^26^.

Detection of the virus in the US improved during 2020 with the ascertainment rate rising from less than 11% in March to nearly 25% in December. Still, the majority of infections remain undocumented, consistent with other estimates^27^. While many of these infections likely present with mild or no symptoms, they remain contagious and support undetected transmission of SARS-CoV-2^5^ making control of the virus very challenging.

Both CFR and IFR declined during 2020. IFR decreased from around 1% in March to about 0.25% in December. Earlier case detection and improved clinical care^28,29^ likely contributed to the decline of both the CFR and IFR during 2020; however, both CFR and IFR are also highly age-dependent with older individuals at substantially greater risk of hospitalization and death^27,30^, so changes to the age distribution of infections over time may have also affected these rates.

Our findings reveal how conditions associated with transmission, case numbers, susceptibility, mortality and control evolved during 2020. Considerable differences in the progression of the pandemic and its epidemiological features manifest in both space and time. In addition, local control efforts appear to have strengthened or slackened in response to increasing or decreasing cases within a locality. This variable responsiveness underscores the need for continued public health messaging emphasizing the maintenance of NPIs while vaccines are distributed and administered.

## Data Availability

Model output and code are posted at https://github.com/SenPei-CU/COVID_US_2020

## Acknowledgements

Mobility data was provided by <u>SafeGraph</u>, a data company that aggregates anonymized location data from numerous applications in order to provide insights about physical places, via the <u>Placekey</u> Community. To enhance privacy, SafeGraph excludes census block group information if fewer than five devices visited an establishment in a month from a given census block group. We also thank Columbia University Mailman School of Public Health for high-performance computing resources. This study was supported by funding from the National Science Foundation (DMS-2027369) and a gift from the Morris-Singer Foundation.

## Author Contributions

SP and JS conceived the study, SP, TKY, SK, MG performed the analysis, SP and JS drafted the manuscript, all authors revised and reviewed the manuscript.

## Data Availability

https://github.com/SenPei-CU/COVID_US_2020

## Supplementary Information

### 1. Transmission model

We use a metapopulation SEIR model to simulate the transmission of COVID-19 in the US at county level. In this model, we explicitly simulate the transmission of documented and undocumented infections, for which separate transmission rates are defined. Further, we assume there is no re-infection of SARS-CoV-2, and that vaccine deployment prior to 31 December 2020 nominally affects population susceptibility.

The model incorporates two types of human mobility across 3,142 US counties – regular daily commuting and diffusive random movement. Information on inter-county commuting is available from the US census survey^1^. During the daytime, commuters travel to counties where they work and mix with the population there; after work, they return home and mix with individuals in their home, residential county. Apart from regular commuting, a fraction of the population in each county, assumed to be proportional to the number of inter-county commuters, travels for purposes other than work. As the population present in each county is different during daytime and nighttime, we model the transmission dynamics of COVID-19 separately for these two time periods. Specifically, we formulate the transmission as a discrete Markov process during both day and night times. The transmission dynamics are depicted by the following equations.

Daytime transmission:

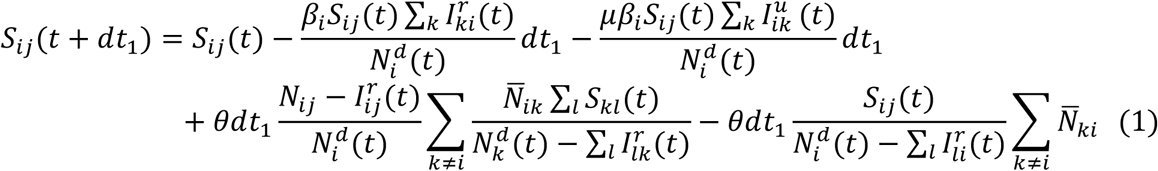

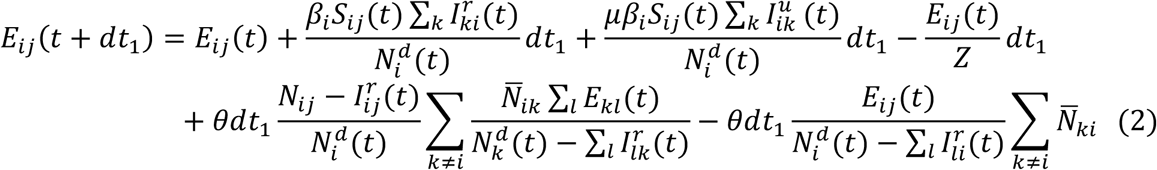

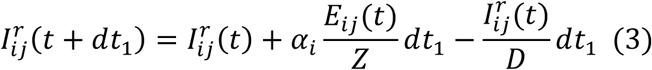

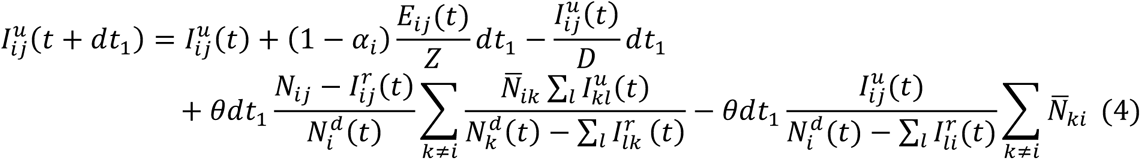

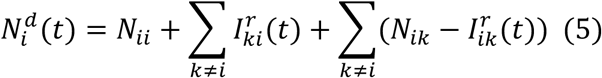

Nighttime transmission:

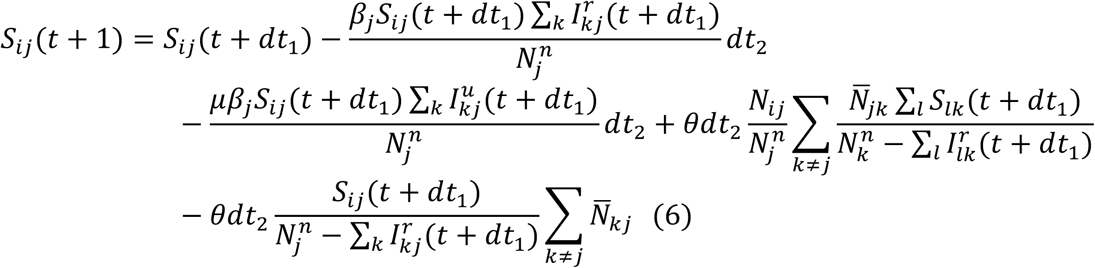

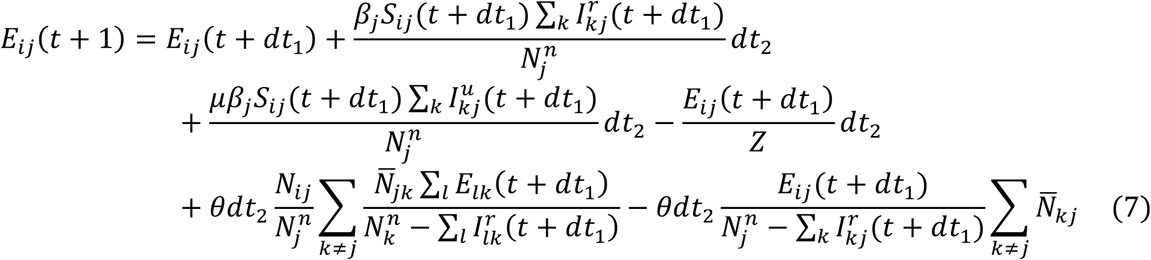

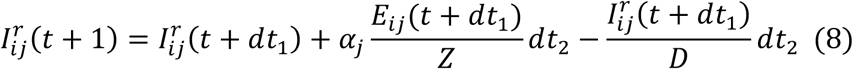

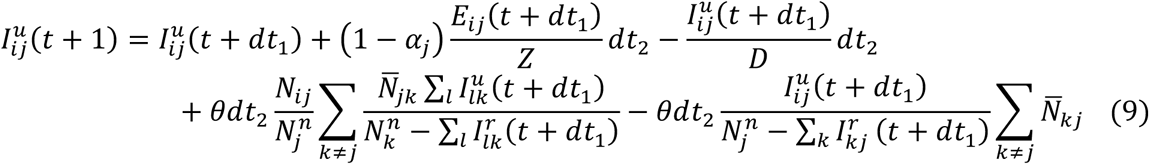

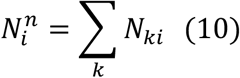

Here, *S*_*ij*_, *E*_*ij*_, 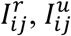, are the susceptible, exposed, documented (reported) infected, undocumented infected and total populations in the subpopulation commuting from county *j* to county *i*(*i*← *j*); *β*_*i*_ is the transmission rate of reported infections in county *i*; *μ* is the relative transmissibility of undocumented infections; *Z* is the average latency period (representing the period from infection to contagiousness); *D* is the average duration of contagiousness; *α*_*i*_ is the fraction of infections documented in county *i*; *θ* is a multiplicative factor adjusting random movement; 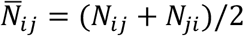 is the average number of commuters between counties *i*and *j*; *dt*_1_ = 1/3 day and *dt*_2_ = 2/3 day are the durations of daytime and nighttime transmission; and 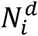 and 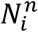 are the daytime and nighttime populations of county *i*. We assume the 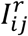 population is immobile and does not participate in human movement. To reflect the spatiotemporal variation of disease transmission rates and reporting, we allowed the transmission rates and ascertainment rates to vary across counties and change over time.

#### Daily work commuting

During the daytime, the population in location *i*, 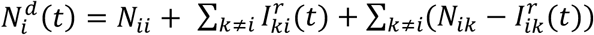, is the sum of individuals who both live and work in location *i*, documented infected individuals who would otherwise commute to other locations *k* (*k* ≠ *i*), and individuals who work in location *i*from other locations *k* (*k* ≠ *i*) but are not documented infections. Within the subpopulation *N*_*ij*_, new infections derive from two processes: contact with documented and undocumented infections in location *i*. For each susceptible individual in *S*_*ij*_(*t*), the chance of contact with documented infections is 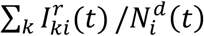, where 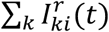 is the total number of documented infections who would commute to all locations *k* but have to stay in location *i*, and the chance of contact with undocumented infections is 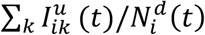, where 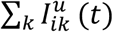 is the total number of undocumented infections in location *i*. Those contacts lead to new infections

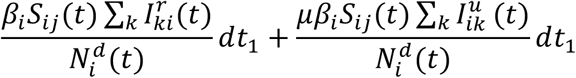

during a period of *dt*_1_ day. Note this term captures the mixing of populations from different locations due to work commuting, and represents intra-county transmission during the daytime in location *i*.

#### Random movement

Apart from work commuting, during the daytime, 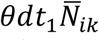 persons, drawn uniformly from the population present in location *k* (*k* ≠ *i*) (except for documented infections) move to location *i*and are randomly redistributed into the subpopulation there. Such population exchange exists for all pairs of locations. For example, for the susceptible population, we first compute the number of susceptible individuals entering into subpopulation *S*_*ij*_(*t*). In other locations *k* (*k* ≠ *i*), the probability a random visitor is susceptible is 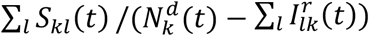, where ∑ _*l*_ *S*_*kl*_ (*t*) is the number of susceptible individuals present in location *k* from all locations *l*, and 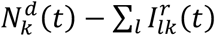 is the total mobile population (i.e., total population minus documented infected population) in location *k*. Therefore, the total susceptible population entering location *i* is 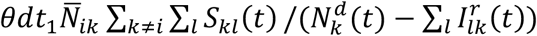. Those individuals are redistributed into subpopulations present in location *i*, where the fraction of people in subpopulation *N*_*ij*_ is 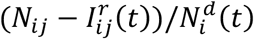. Finally, the number of susceptible individuals entering *S*_*ij*_(*t*) is

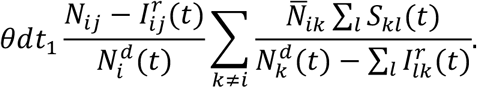

We then compute the number of susceptible individuals leaving *S*_*ij*_(*t*). The total number of individuals leaving location *i* is 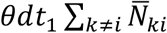; the fraction of susceptible people from *N*_*ij*_ is 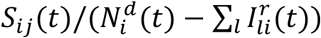. As a result, the number of susceptible persons leaving *S*_*ij*_(*t*) is

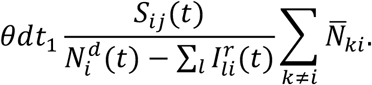

Population exchange for other compartments can be computed similarly. Note there is no random movement in Eq. (3) as we assume documented infections, 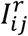, are immobile. We can write Eqs. (6-10) for nighttime transmission similarly.

Prior to March 1, 2020, we used the commuting data from the US census survey to prescribe the inter-county movement in the transmission model. After March 1, the census survey data are no longer representative due to changes in mobility behavior following implementation of non-pharmaceutical interventions. We therefore used estimates of the reduction of inter-county visitors to points of interest (POI) (e.g., restaurants, stores, etc.) from SafeGraph^2^ to account for the change of inter-county movement on a county-by-county basis. For instance, if the number of inter-county visitors in a county was reduced by 10% on a given day relative to the baseline on March 1, the number of commuters and random visitors to this county would be reduced by 10% accordingly.

The transmission model generates daily confirmed cases for each county. To account for reporting delays, we mapped simulated documented infections to confirmed cases using a separate observational delay model. In this delay model, we account for the time interval between a person transitioning from latent to contagious (i.e., *E* → *I*^*r*^) and confirmation of that individual infection. To estimate this delay period, *T*_*r*_, we examined a U.S. line list data record consisting of 13.4 million confirmed cases until December 31, 2020^3^. We used a gamma distribution to fit the time-to-event distribution of the interval (in days) from symptom onset to case confirmation for each month in 2020. We modeled *T*_*r*_ by adding another 2.5 days to the mean periods of the obtained gamma distributions, as symptom onset is estimated to lag the onset of contagiousness^4^. The gamma distributions of *T*_*r*_ from April to December are provided in Table S1.

### 2. Model calibration

We calibrated the transmission model against county-level incidence data reported from 21 February 2020 to 31 December 2020, available at Johns Hopkins University coronavirus resource center^5^. Model parameters were estimated using a sequential data assimilation method – the ensemble adjustment Kalman filter (EAKF)^6^, which is applicable to high-dimensional metapopulation models and has been successfully used to infer epidemiological parameters for a range of infectious diseases^7–11^. To represent the state-space distribution (including both parameters and variables), the EAKF maintains an ensemble of system state vectors acting as samples from the distribution. In particular, the EAKF assumes a Gaussian distribution of both the prior and likelihood and adjusts the prior distribution to a posterior using Bayes’ rule: posterior ∝ prior × likelihood. For the observed variables (i.e., daily incidence), ensemble members are updated deterministically such that the higher moments of the prior distribution are preserved in the posterior. Unobserved variables and parameters are updated based on their covariability with the observed variable, which can be computed directly from the ensemble. Further details on the EAKF scheme can be found in Anderson^6^.

We derived the estimate of model parameters by coupling the EAKF algorithm with the disease transmission model. To further reduce the number of unknown parameters in this high-dimensional transmission model, we fixed disease-related parameters (*Z, D*, and *μ*) and the mobility factor (*θ*) as estimated using case data through 13 March 2020^12^. Specifically, these parameters were randomly drawn from the posterior distributions: *Z* = 3.59 (95% CI: 3.28 – 3.99), *D* = 3.56 (3.21 – 3.83), *μ* = 0.64 (0.56 – 0.70), and *θ* = 0.15 (0.12 – 0.17). We performed EAKF inference each day using case data in 3,142 US counties to estimate the ascertainment rate *α*_*i*_ and transmission rate *β*_*i*_. To account for the reporting delay of confirmed cases, at each daily model update, we integrated the model forward 9 days using the prior model state and used incidence 9 days ahead (i.e., roughly the modes of gamma distributions for delays) to constrain current model variables and parameters.

To initialize the model-inference system, we seeded exposed individuals (*E*) and undocumented infections (*I*^*u*^) in counties with at least five confirmed case. Specifically, we randomly drew *E* and *I*^*u*^ from uniform distributions [0, 12*C*] and [0, 10 *C*] 9 days before the first date with more than five reported cases, *T*_*0*_, where *C* is the total number of reported cases between day *T*_*0*_ and *T*_*0*_ + 4. This setting provides a broad seeding range for US counties. The prior ascertainment rates were drawn from a distribution with a median value *α* = 0.080 (0.069 – 0.093), estimated using case data prior to 13 March 2020. The prior transmission rates were scaled on the basis of the local population density: *β*_*i*_ = *β*_*0*_ × log_1*0*_ *PD*_*i*_ /median(log_1*0*_ *PD*), where *PD*_*i*_ is the population density in county *i*, median(log_1*0*_ *PD*) is the median value of log-transformed population density among all counties, and *β*_*0*_ = 0.95 (0.84 – 1.06) is the baseline transmission rate estimated before 13 March 2020. For counties that reported less than 20 cumulative cases as of 15 March 2020, we reduced the prior transmission rate by half to reflect the impact of non-pharmaceutical interventions implemented after the announcement of national emergency. We performed the inference using 100 ensemble members.

### 3. Metropolitan areas

In this study, we report the transmission dynamics in and around five major US cities: New York, Chicago, Los Angeles, Phoenix, and Miami. Characteristics of SARS-CoV-2 transmission (e.g., ascertainment rate, CFR, and IFR) in these metropolitan areas were aggregated from county-level estimates. Note that in some cases the metropolitan areas as defined here differ from the formal metropolitan statistical areas delineated by the United States Office of Management and Budget. The counties we include in the metropolitan areas for this analysis are:

1. New York: Kings County NY, Queens County NY, New York County NY, Bronx County NY, Richmond County NY, Westchester County NY, Bergen County NJ, Hudson County NJ, Passaic County NJ, Putnam County NY, and Rockland County NY
2. Chicago: Cook County IL, DuPage County IL, Kane County IL, McHenry County IL, and Will County IL
3. Los Angeles: Los Angeles County CA and Orange County CA
4. Phoenix: Maricopa County AZ, Pinal County AZ, and Gila County AZ
5. Miami: Miami-Dade County FL, Broward County FL, and Palm Beach County FL

### 4. Seroprevalence surveys

We validated the estimated cumulative infections against the proportion of SARS-CoV-2 seropositive individuals from two large-scale serological surveys in the US: 1) surveys in 10 sites until August 2020^13,14^, and 2) state-level surveys from August 2020 to November 2020^15,16^. We primarily focused on the 10-site survey as samples were collected in more consistent and specific locations.

The serological surveys give an indication of the proportion of individuals previously infected by SARS-CoV-2 and in principle should be similar to the proportion of cumulative infections estimated by the model. However, the seroprevalence rates likely underestimate the fraction of a population previously infected with SARS-COV-2 due to antibody waning. Specifically, antibody titers in recovered individuals decline over time, and seroreversion in the months following adaptive immune response is common^17^. Note, such decline in antibody titers does not necessarily preclude protection from repeat infections mediated by other components of the adaptive immune response.

We adjusted the reported seroprevalence by first correcting for errors in serological testing. The assay used to detect antibodies against SARS-CoV-2 reports 96% (95% CI: 98.3 – 99.9%) sensitivity and 99.3% (98.3 – 99.9%) specificity^13^. The seroprevalence adjusted for testing sensitivity and specificity is *p* = (*p*_*report*_ + *specificity* – 1)/(*sensitivity* + *specificity* − 1), where *p*_*report*_ is the reported seroprevalence^18^.

We then adapted the method of Buss et al. to quantify antibody waning (seroreversion) and to estimate an adjusted seroprevalence that is inclusive of individuals who have seroreverted^19^. This method has been used to estimate the percentage of the population infected with SARS-CoV-2 in Manaus, Brazil during a largely unmitigated outbreak^19^. In order to estimate the antibody waning rate, we used the seroprevalence data for New York City from the 10-site study. As New York City was the epicenter in the US during the early phase of the COVID-19 pandemic, the effect of seroreversion is expected to be more apparent there than for other locations.

We assume the probability of seroreversion for an infected patient decays exponentially with time, and define the monthly attenuation as *w* ∈ [0,1]. The probability of a recovered patient seroreverting after *m* months is (1 − *w*)*w*^*m*^/*w*. Denote ***r*** = [*r*(*t*_1_), …, *r*(*t*_*n*_)]^*T*^ and ***d*** =[*p*(*t*_1_), *p*(*t*_2_) − *p*(*t*_1_), …, *p*(*t*_*n*_) − *p*(*t*_*n*_ − 1)]^*T*^, where *r*(*t*_*i*_) is the number of new recoveries per capita at time *t*_*i*_, and *p*(*t*_*i*_) is the measured seroprevalence. Here we define *t*_*i*_ as the mid-point of the sampling period for the *i*th serological survey. We have ***d*** = ***Ar*** (see details in Buss et al.^18^), where the *n* × *n* matrix ***A*** has entries

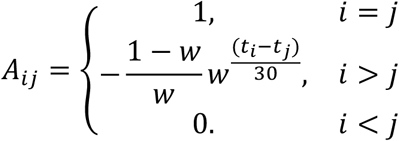

The triangular matrix ***A*** is always invertible as the diagonal elements are all ones. The new recoveries ***r*** can be computed by ***r*** = ***A***^-1^***d***.

To estimate *w*, we first sampled *p*(*t*_*i*_) (*i*= 1, …, 6) from a uniform distribution [*p*_*lower*_(*t*_*i*_), *p*_*upper*_(*t*_*i*_)], where *p*_*lower*_(*t*_*i*_) and *p*_*upper*_(*t*_*i*_) are the observed lower and upper bounds of seroprevalence adjusted for testing characteristics. We then searched a range of linearly spaced numbers *w* from 0.001 to 0.999 and computed ***r*** for each *w* using the sampled seroprevalence data from New York City. As New York City reported few cases in July, we estimated *w* as the value that minimizes the number of new recoveries in the last two sampling periods (i.e., *r*(*t*_5_) + *r*(*t*_6_), July 7 to 11 and July 27 to 30 July) under the constraint *r*(*t*_*i*_) ≥ 0 for all *t*_*i*_. To account for uncertainty in observed seroprevalence, we independently drew 100 samples of ***d***, and selected the *w* value that generated the lowest new recoveries in July among all ***d*** samples. This process was repeated 1,000 times to obtain the distribution of monthly attenuation: *w* = 0.90 (95% CI: 0.86 – 0.95). The adjusted seroprevalence in New York City is the cumulative number of recoveries per capita computed using the estimated parameter *w*. For other locations, we used the same parameter *w* to correct the prevalence if it generates non-negative *r*(*t*_*i*_) for all *t*_*i*_. Otherwise, we chose the parameter closest to *w* in New York City that produces non negative *r*(*t*_*i*_)^18^. The adjusted and reported seroprevalence for the 10 sites are shown in Fig. S2. To match the estimated cumulative infection and seroprevalence in one location, we assumed seroconversion takes an average of 10 days (e.g., from infection acquisition to generation of detectable antibody). A 14-day seroconversion period was also tested and results remained similar.

We applied the same method to adjust state-level seroprevalence reported from August to November 2020 (Fig. S3). However, as seroprevalence data were not available prior to August, the adjustment cannot account for seroreversion before August 2020. As a result, the adjusted seroprevalence in August is an underestimate. In addition, because the sample size is small relative to the population of each state and the samples may not be representative of general US population, anomalies may appear in the reported seroprevalence. For instance, the survey in North Dakota estimated 7.3% seroprevalence between 29 July and 12 August; however, it dropped to 0.6% between 12 August and 25 August. To exclude any severely biased seroprevalence data, we assumed the monthly attenuation rate to be no higher than 15%. Observations that indicated faster antibody waning (e.g., North Dakota) were excluded from the analysis. In total, seroprevalence data in 17 states were used (Fig. S3a). Despite these limitations, our inferred cumulative infected percentages are well matched to adjusted seroprevalence at state level (Fig. S4a, Pearson *r* = 0.76, mean absolute error (MAE) = 3.61%). Note that the adjusted seroprevalence data in August (blue dots) are generally lower than model estimates, as seroreversion prior to August was not considered in the adjustment. For later surveys (yellow nodes), this systematic bias is less severe. We further performed the same analysis using a maximum monthly attenuation rate of 25% (Fig. S3b and Fig. S4b, Pearson *r* = 0.72, mean absolute error (MAE) = 4.57%).

### 5. Characteristics of COVID-19

The estimated monthly infections (both documented and undocumented) in the US and five metropolitan areas are reported in Fig. S5. The prevalence of contagious infections in the community is estimated as the percentage of active infectious cases (*I*^*r*^ + *I*^*u*^) among the general population. We provide the daily confirmed cases and estimated prevalence of contagious infections in the US and five metropolitan areas in Fig. S6.

We estimated the CFR and IFR for patients infected each day. Specifically, we define the number of infections (both documented and undocumented) on day *t* as 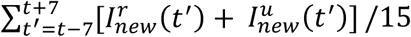, where 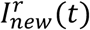 and 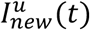 are the posterior newly infected reported and undocumented patients on day *t*. We use the average of new infections during a 15-day time window to smooth the variation of reporting within a week. A proportion of total infections were tested and reported as confirmed cases after a lag between infection acquisition and laboratory confirmation. Denote *T*_*r*_ as the mean reporting delay on day *t* (see Table S1). We estimated the number of confirmed cases who were infected on day *t* as 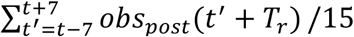, where *obs*_*post*_(*t*) is the posterior confirmed cases on day *t*. Note here we shifted the computation time window forward for *T*_*r*_ days to account for reporting delays. Based on observations of confirmed COVID-19-related deaths in New York City^20^, the mean time from confirmation to death is *T*_*d*_ = 9 days. We therefore estimated the number of deaths among patients infected on day *t* as 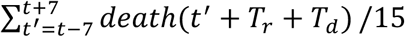, where *death*(*t*) is the number of reported COVID-19-related deaths on day *t*. The CFR is computed as 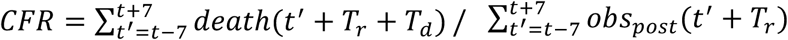, and the IFR is computed as 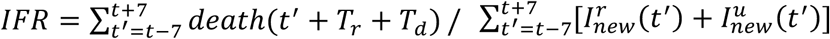. As death data for individuals infected in late December were not available when we performed the study, we limited analysis on CFR and IFR until 1 December 2020.

As the mortality rate has large variations across different age groups, the overall IFR at a given time depends on the age structure of infections. The fraction of confirmed infections for each age group in HHS region 4 (Alabama, Florida, Georgia, Kentucky, Mississippi, North Carolina, South Carolina, and Tennessee) reported by the CDC^21^ is shown in Fig. S7.

## 6. Quantifying *R*_*t*_ change in response to local COVID-19 infections

We use the time-varying reproduction number, *R*_*t*_ = *βD*[*α* + *μ*(1 − *α*)], to quantify the local transmission rate of COVID-19. Based on the national confirmed cases in the US, we define the spring, summer and fall/winter waves as the following periods: 21 January 2020 to 31 May 2020, 1 June 2020 to 15 September 2020, and 16 September 2020 to 31 December 2020. For each wave, we selected the time intervals in each county with increasing and decreasing local infections. Specifically, if the weekly confirmed cases (per 100,000 people) in a county increase (or decrease) for at least two consecutive weeks, this period is defined as an interval with increasing (or decreasing) local transmission for the county. To remove the counties with low activity, we discarded those that never reported greater than 15 daily cases per 100,000 population during a wave. For counties with increasing local infections, we included 352, 492 and 594 counties in the analysis for the spring, summer, and fall/winter waves. For counties with decreasing local infections, 114, 356 and 288 counties were included in the analysis for the spring, summer, and fall/winter waves. Analyses using other threshold values (25 and 40) yield similar results (Figs. S8-S9). We computed the weekly reproduction number *R*_*t*_ in counties with increasing (or decreasing) infections and fitted the estimate *R*_*t*_ to EPI week using a linear function for each wave (EPI week is the epidemiological week used by the US CDC). The response to local transmission is quantified by the slope of the linear fit, which is interpreted as the weekly change of *R*_*t*_.

---

**Table S1.** Monthly gamma distributions for the reporting delay *T*_*r*_.

| Months | Shape parameter $a$ | Scale parameter $b$ | Mean (days) |
| --- | --- | --- | --- |
| $\leq 4$ | 4.27 | 3.25 | 13.9 |
| 5 | 4.06 | 3.23 | 13.1 |
| 6 | 4.33 | 3.03 | 13.1 |
| 7 | 4.18 | 3.11 | 13.0 |
| 8 | 3.91 | 3.09 | 12.1 |
| 9 | 3.41 | 3.31 | 11.3 |
| 10 | 3.10 | 3.59 | 11.2 |
| 11 | 3.47 | 3.55 | 12.3 |
| 12 | 3.10 | 3.99 | 12.4 |

**Fig. S1.**
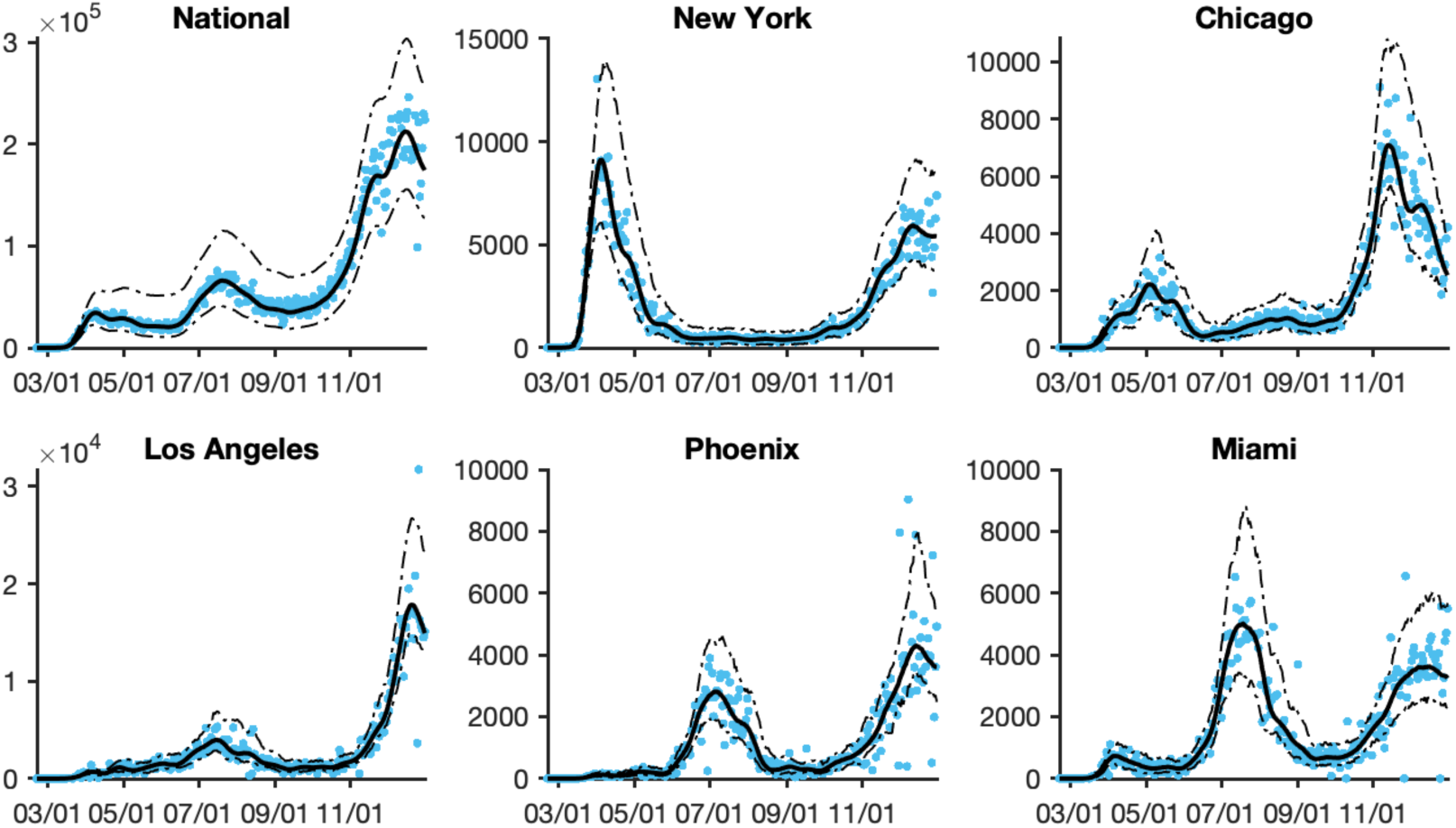
Model fitting to daily case numbers (blue dots) in the US and five metropolitan areas. Solid and dashed lines show the median estimate and 95% CIs respectively.

**Fig. S2.**
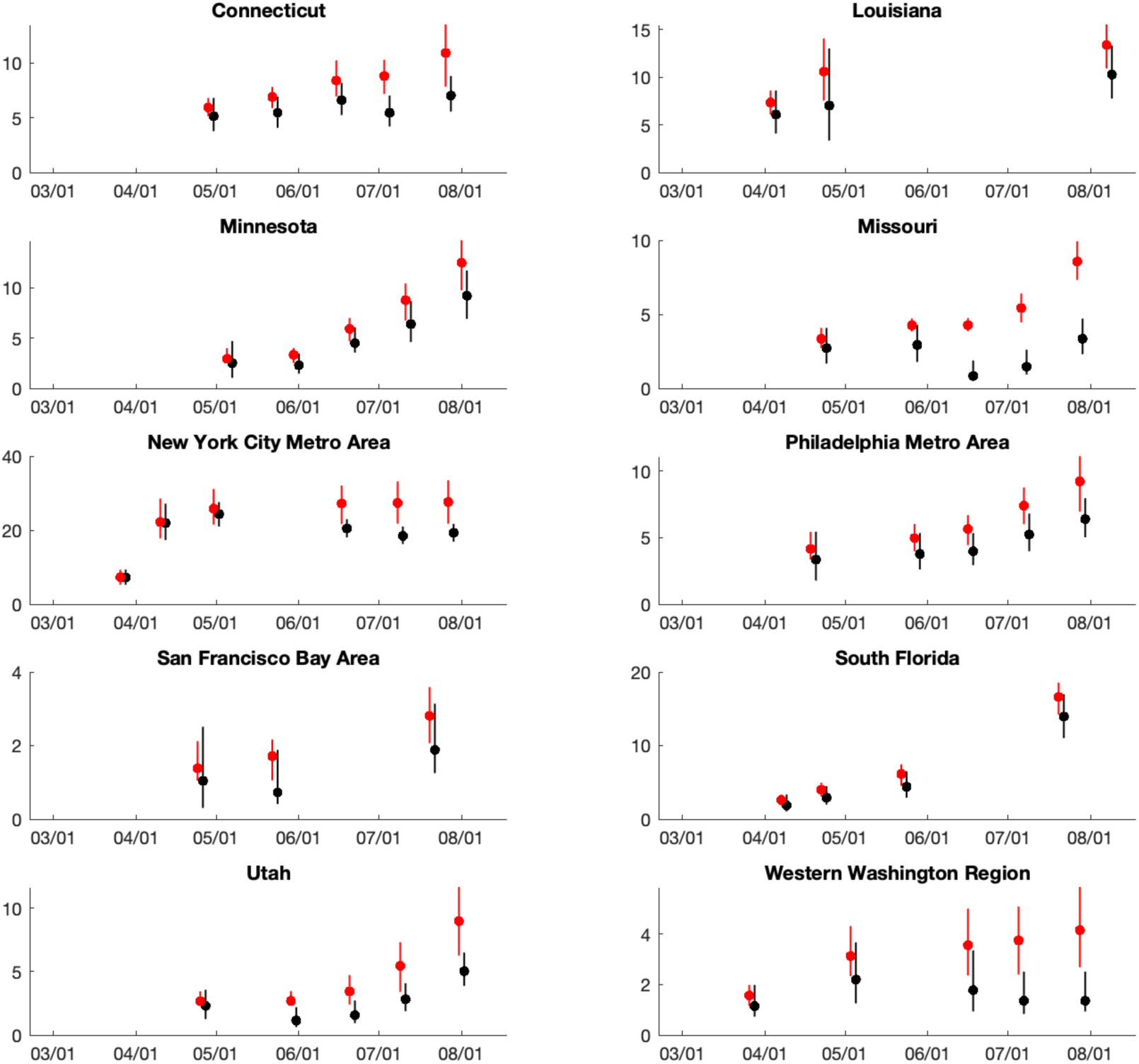
The reported (black) and adjusted (red) seroprevalence in the 10-site study. Dots and whiskers show the median and 95% CIs respectively.

**Fig. S3.**
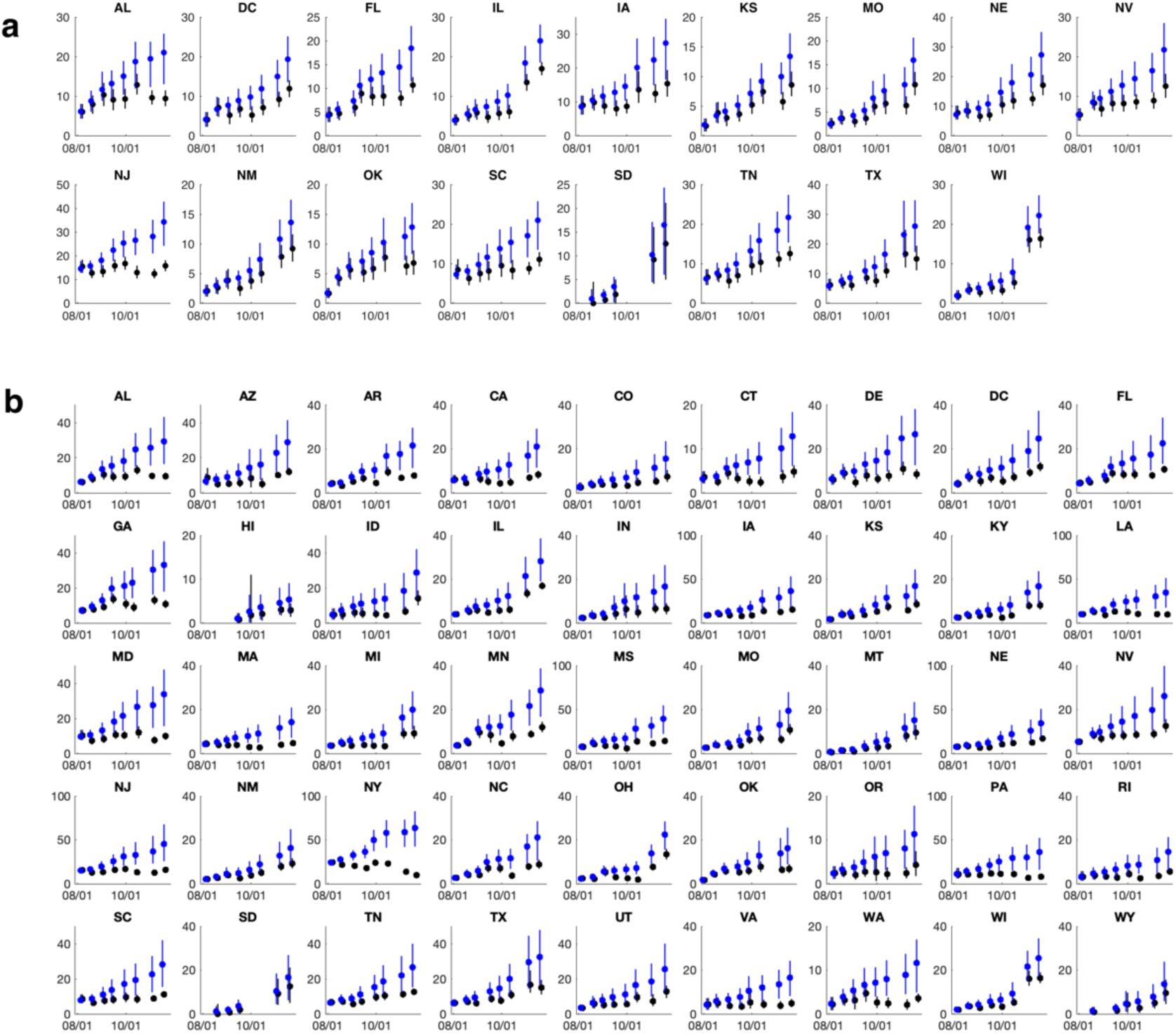
The reported (black) and adjusted (blue) seroprevalence for the state-level serological survey. Dots and whiskers show the median and 95% CIs. (a) and (b) show the results obtained using a maximum monthly attenuation rate of 15% and 25%, respectively.

**Fig. S4.**
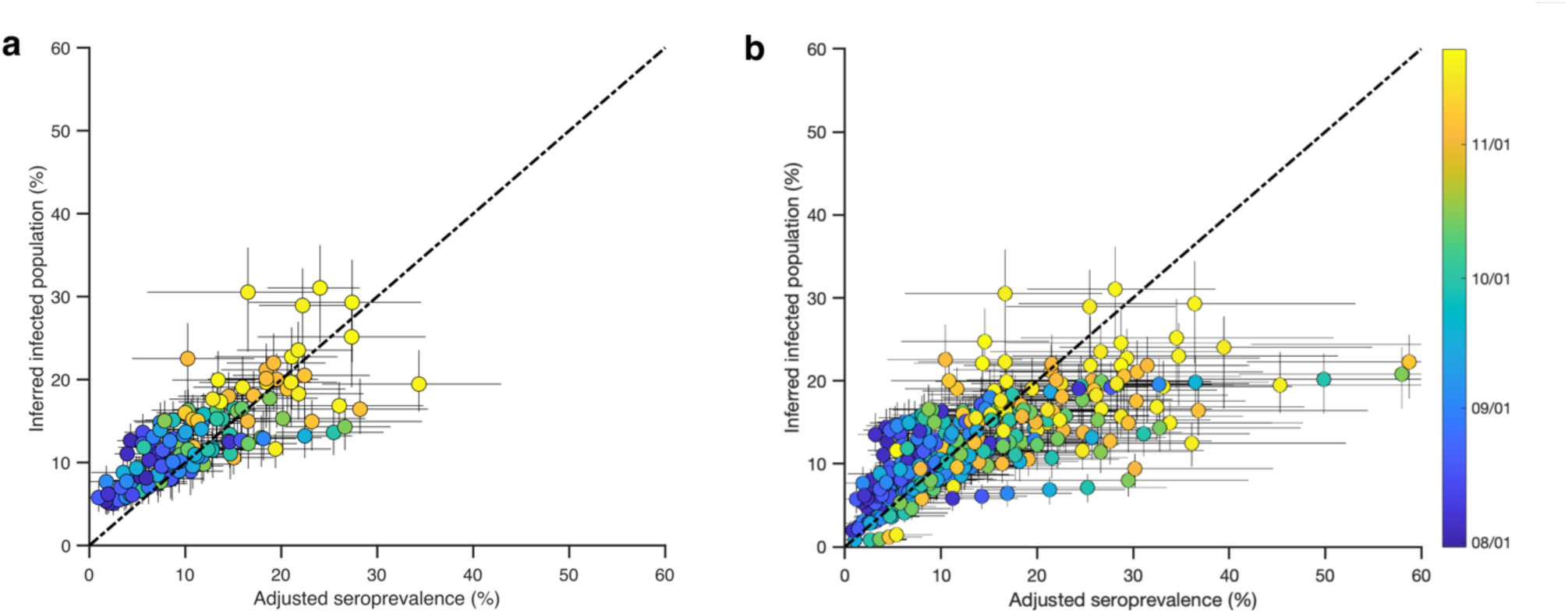
Comparison between the inferred percentage of cumulative infections and seroprevalence at the state level adjusted for antibody waning. Whiskers show 95% CIs and color indicates the sample collection date for each location. Seroprevalence data adjusted using a maximum monthly attenuation rate of 15% (a) and 25% (b) are included in the analysis.

**Fig. S5.**
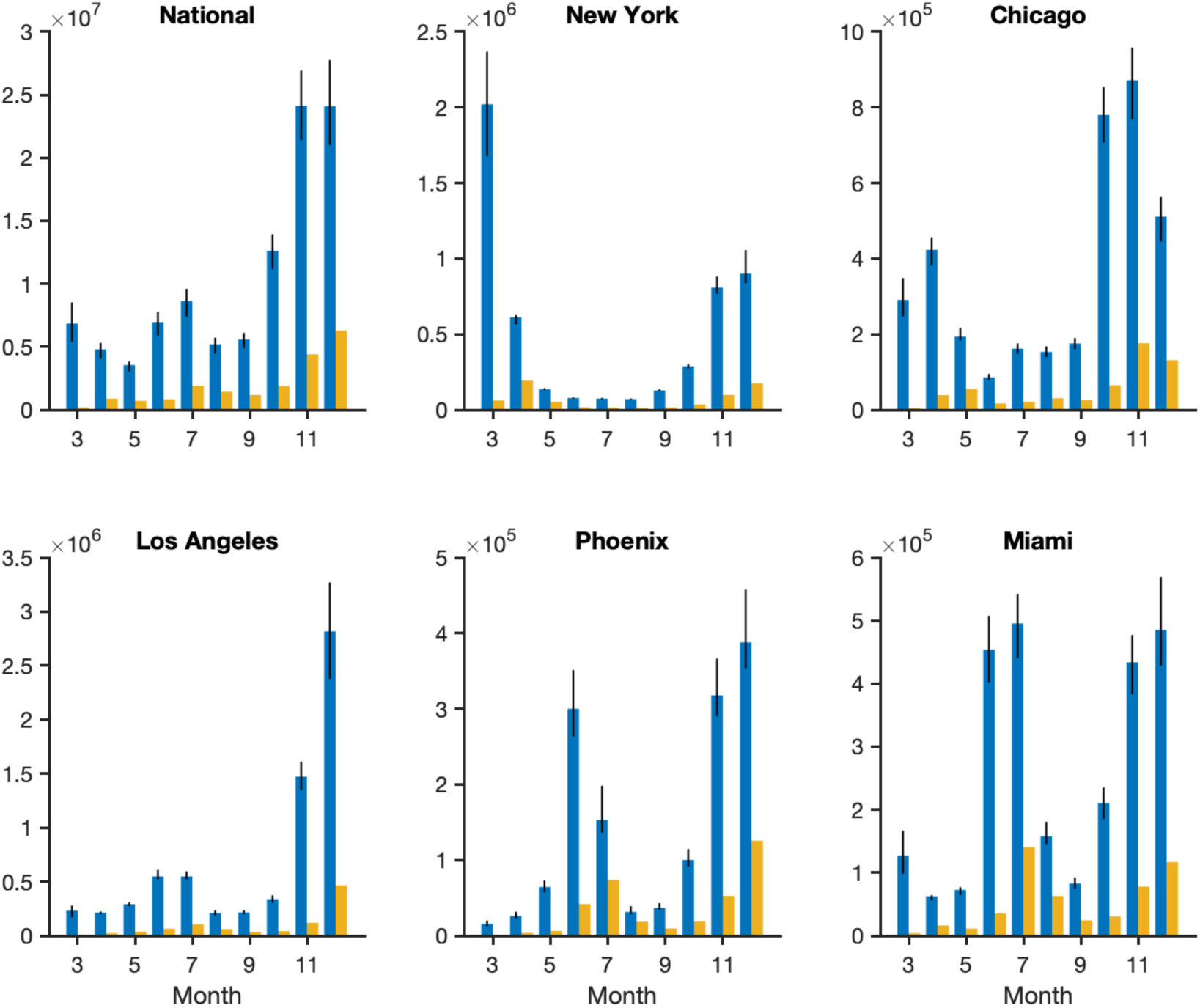
Estimated monthly total infections (blue bars, whiskers show 95% CIs) and confirmed cases (orange bars) in the US and five metropolitan areas.

**Fig. S6.**
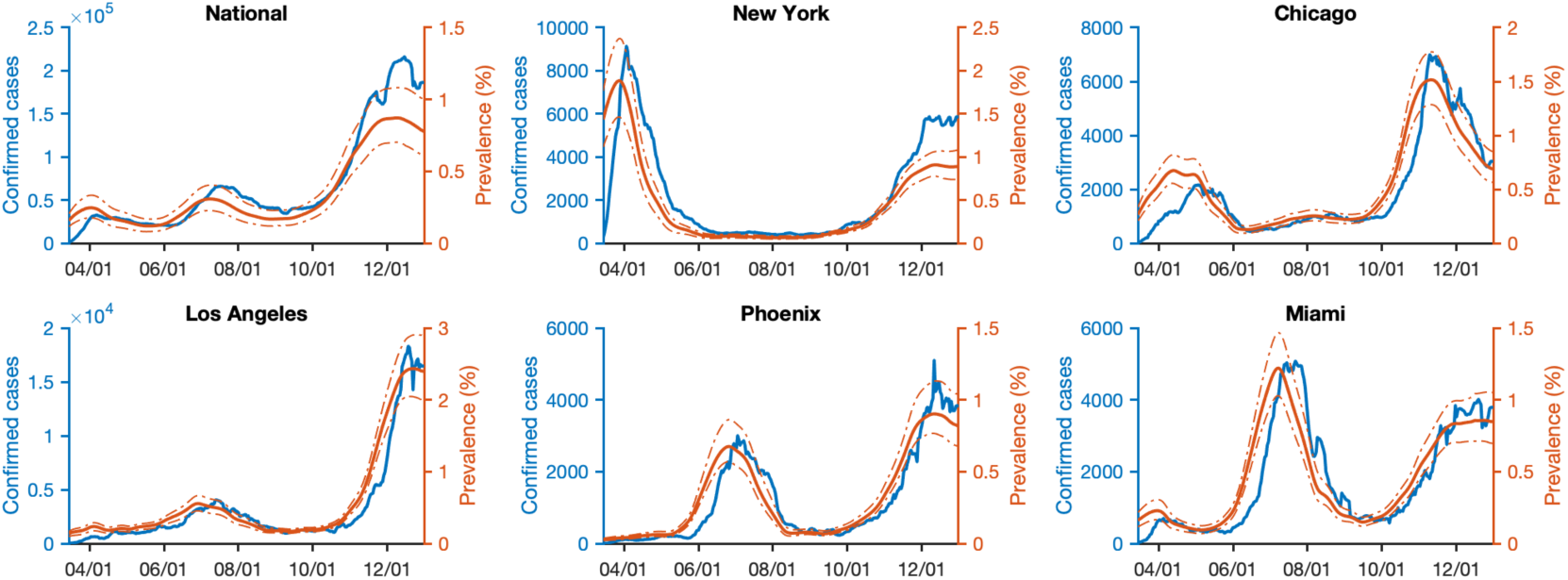
Daily confirmed cases (blue line, 7-day moving average) and estimated prevalence of contagious infections (red line, median and 95% CIs) in the US and five metropolitan areas.

**Fig. S7.**
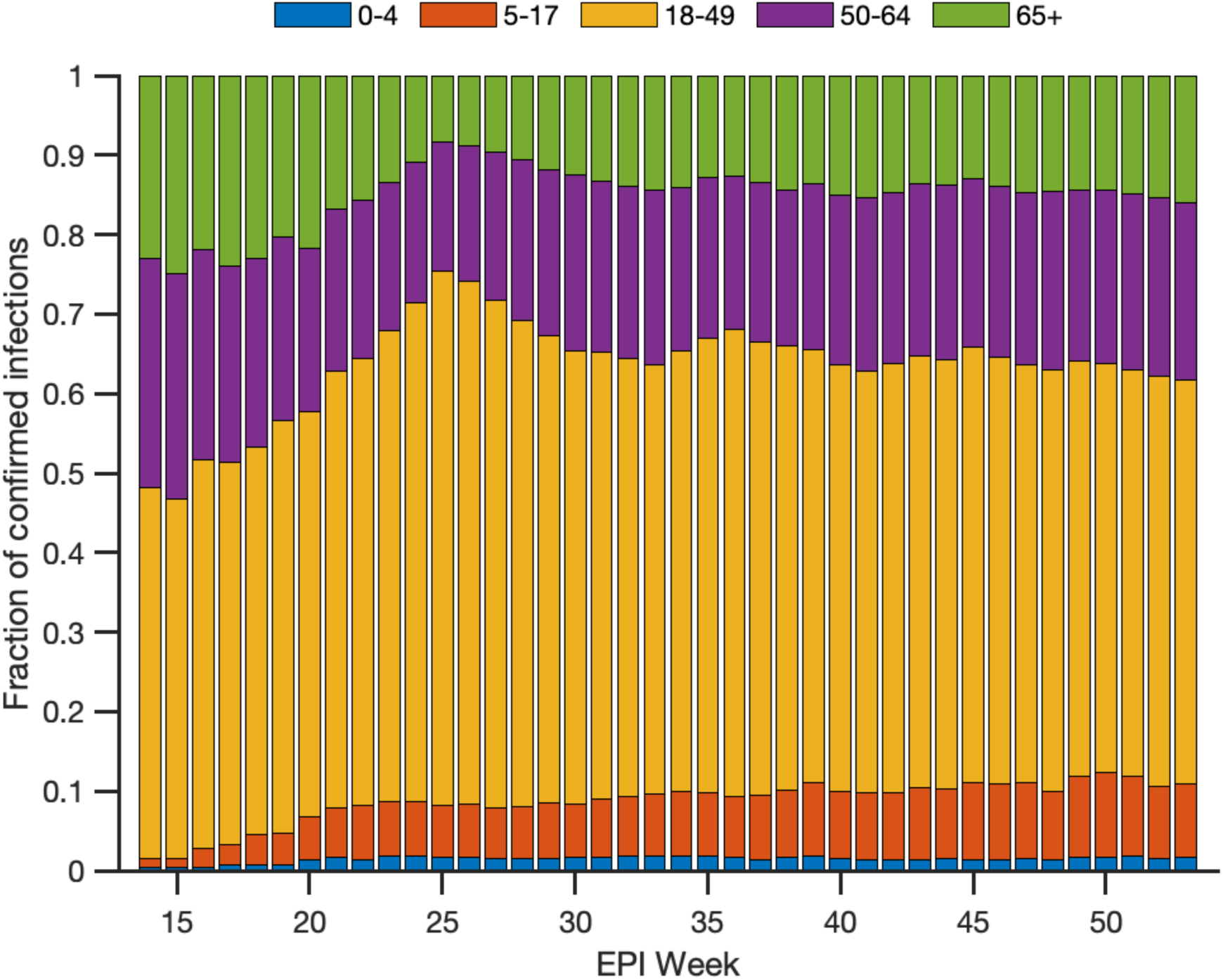
Age distribution of confirmed infections in US Health and Human Services Region 4 (Alabama, Florida, Georgia, Kentucky, Mississippi, North Carolina, South Carolina, and Tennessee).

**Fig. S8.**
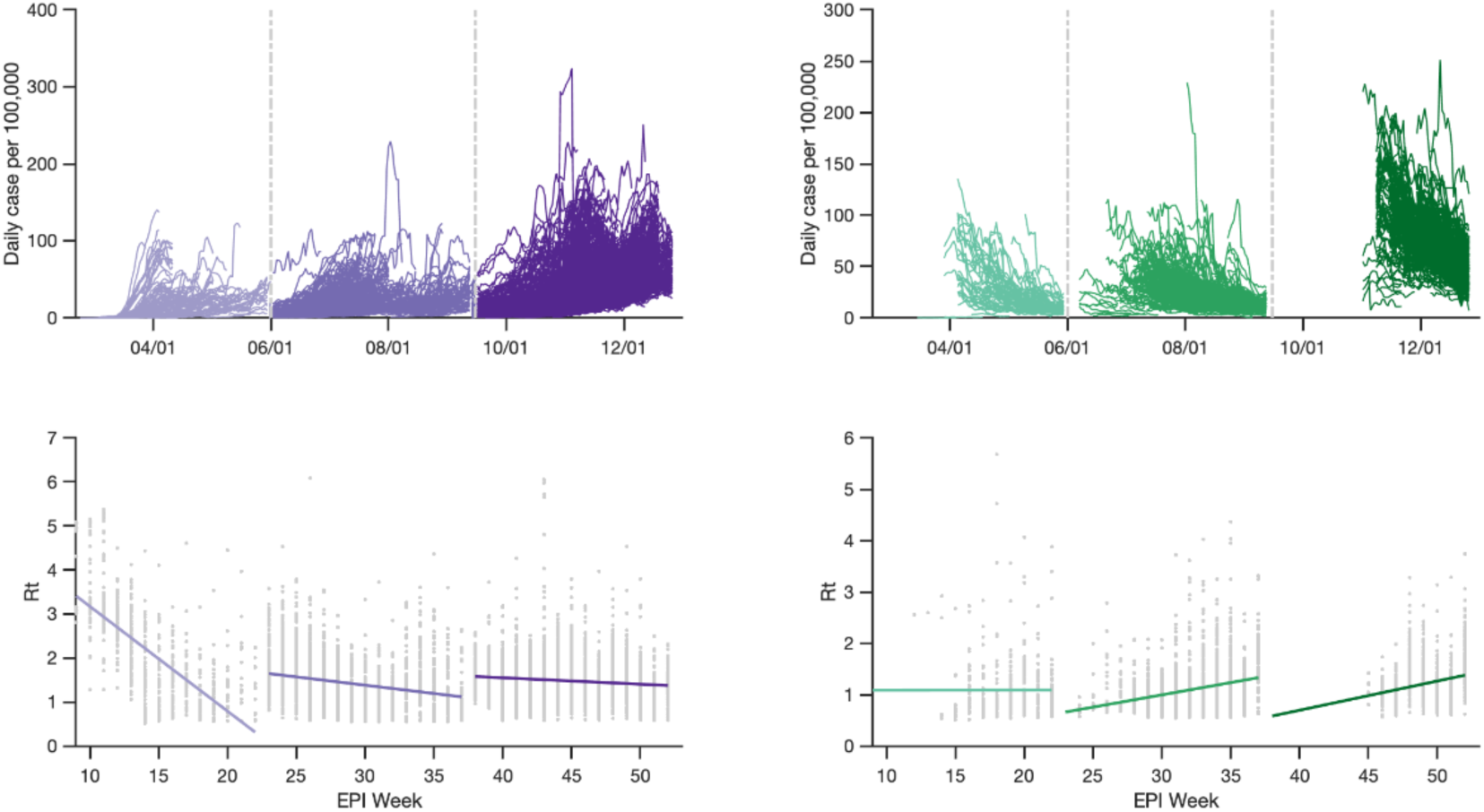
Response to growth and decline of local infections. Curves with the maximum case per 100,000 people below 25 are removed in this analysis.

**Fig. S9.**
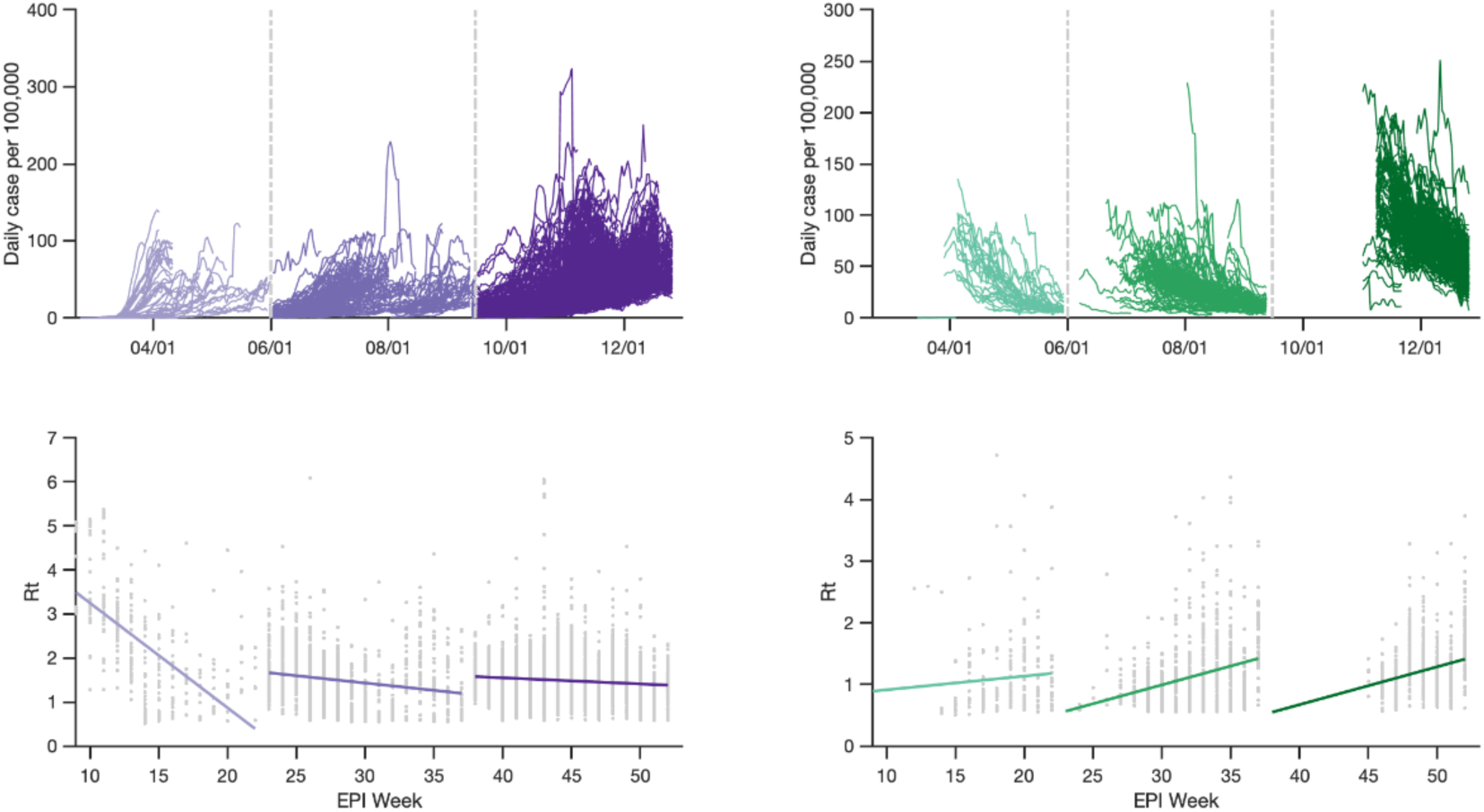
Response to growth and decline of local infections. Curves with the maximum case per 100,000 people below 40 are removed in this analysis.

## References

1. Li, Q., et al. Early transmission dynamics in Wuhan, China of novel coronavirus-infected pneumonia. New Engl. J. Med. 382, 1199–1207 (2020).

2. Pekar, J., Worobey, M., Moshiri, N., Scheffler, K. & Wertheim, J. O. Timing the SARS-CoV-2 index case in Hubei province. bioRxiv, 392126 (2020).

3. Ng, K.W., et al. Preexisting and de novo humoral immunity to SARS-CoV-2 in humans. Science 370, 1339–1343 (2020).

4. Wu, J. T., Leung, K. & Leung, G. M. Nowcasting and forecasting the potential domestic and international spread of the 2019-nCoV outbreak originating in Wuhan, China: a modelling study. Lancet 395, 689–697 (2020).

5. Li, R., et al. Substantial undocumented infection facilitates the rapid dissemination of novel coronavirus (SARS-CoV-2). Science 368, 489–493 (2020).

6. Arons, M. M., et al. Presymptomatic SARS-CoV-2 infections and transmission in a skilled nursing facility. New Engl. J. Med. 382, 2081–2090 (2020).

7. He, X., et al. Temporal dynamics in viral shedding and transmissibility of COVID-19. Nature Med. 26, 672–675 (2020).

8. World Health Organization, WHO Coronavirus Disease (COVID-19) Dashboard. https://covid19.who.int (2021).

9. U.S. Food and Drug Administration, Pfizer-BioNTech COVID-19 Vaccine. https://www.fda.gov/emergency-preparedness-and-response/coronavirus-disease-2019-covid-19/pfizer-biontech-covid-19-vaccine (2020).

10. U.S. Food and Drug Administration, Moderna COVID-19 Vaccine. https://www.fda.gov/emergency-preparedness-and-response/coronavirus-disease-2019-covid-19/moderna-covid-19-vaccine (2020).

11. Pei, S., Kandula, S. & Shaman, J. Differential effects of intervention timing on COVID-19 spread in the United States. Science Adv. 6, eabd6370 (2020).

12. Yamana, T., Pei, S. & Shaman, J. Projection of COVID-19 cases and deaths in the US as individual states re-open, May 4, 2020. medRxiv, 90670 (2020).

13. U.S. Centers for Disease Control and Prevention, CDC COVID Data Tracker, https://covid.cdc.gov/covid-data-tracker/?CDC_AA_refVal=https%3A%2F%2Fwww.cdc.gov%2Fcoronavirus%2F2019-ncov%2Fcases-updates%2Fcommercial-labs-interactive-serology-dashboard.html#testing_positivity7day (2020).

14. Buss, L. F. et al.. Three-quarters attack rate of SARS-CoV-2 in the Brazilian Amazon during a largely unmitigated epidemic. Science 371, 288–292 (2021).

15. Shioda, K. et al.. Estimating the cumulative incidence of SARS-CoV-2 infection and the infection fatality ratio in light of waning antibodies. medRxiv, 231266 (2020).

16. U.S. Centers for Disease Control and Prevention, Disease Burden of Influenza. https://www.cdc.gov/flu/about/burden/index.html?CDC_AA_refVal=https%3A%2F%2Fwww.cdc.gov%2Fflu%2Fabout%2Fdisease%2Fburden.htm&wdLOR=c30DA53FC-A68A-D745-BACF-4B86F2B93082 (2021).

17. Riley, S., et al. Epidemiological characteristics of 2009 (H1N1) pandemic influenza based on paired sera from a longitudinal community cohort study. PLoS Med. 8, e1000442 (2011).

18. Ma, Y., Pei, S., Shaman, J., Dubrow, R. & Chen K. Role of air temperature and humidity in the transmission of SARS-CoV-2 in the United States. medRxiv, 231472 (2020).

19. COVID Analysis and Mapping of Policies, https://covidamp.org/policymaps (2021).

20. To, K. K.-W., et al. Coronavirus disease 2019 (COVID-19) re-infection by a phylogenetically distinct severe acute respiratory syndrome coronavirus 2 strain confirmed by whole genome sequencing. Clin. Inf. Dis. ciaa1275 (2020).

21. Tillett, R. L., et al. Genomic evidence for reinfection with SARS-CoV-2: a case study. Lancet Inf. Dis. 21, 52–58 (2021).

22. Self, W. H. et al.. Decline in SARS-CoV-2 antibodies after mild infection among frontline health care personnel in a multistate hospital network - 12 states, April-August 2020. Morb. Mort. Weekly Rep. 69, 1762–1766 (2020).

23. Choe, P. G., et al. Waning antibody responses in asymptomatic and symptomatic SARS-CoV-2 infection. Emerg. Inf. Dis. 27, 327–329 (2021).

24. Fiorentini, S., et al. First detection of SARS-CoV-2 spike protein N501 mutation in Italy in August, 2020. Lancet Inf. Dis. 21, s1473–3099 (2021).

25. Rambaut, A., et al. Preliminary genomic characterization of an emergent SARS-CoV-2 lineage in the UK defined by a novel set of spike mutations. https://virological.org/t/preliminary-genomic-characterisation-of-an-emergent-sars-cov-2-lineage-in-the-uk-defined-by-a-novel-set-of-spike-mutations/563 (2020).

26. Shaman, J. & Galanti, M. Will SARS-CoV-2 become endemic? Science 370, 527–529 (2020).

27. Reese, H. et al. Estimated incidence of COVID-19 illness and hospitalization – United States, February-September, 2020. Clin. Inf. Dis. ciaa1780 (2020).

28. Hallifax, R. J. et al. Successful awake proning is associated with improved clinical outcomes in patients with COVID-19: single-centre high-dependency unit experience. BMJ Open 7, e000678 (2020).

29. Beigel, J. H. et al. Remdesivir for the treatment of COVID-19 – final report. New Engl. J. Med. 383, 1813–1826 (2020).

30. Yang, W. et al. Estimating the infection fatality risk of SARS-CoV-2 in New York City during the spring 2020 pandemic wave: a model-based analysis Lancet Inf. Dis. 21, 203–212 (2021).

## References

1. Bureau, U. C. County to county commuting data. The United States Census Bureau https://www.census.gov/topics/employment/commuting.html (2013).

2. SafeGraph | POI Data, Business Listings, & Foot-Traffic Data. https://www.safegraph.com/.

3. Calgary, O. COVID-19 Case Surveillance Public Use Data | Data | Centers for Disease Control and Prevention. https://data.cdc.gov/Case-Surveillance/COVID-19-Case-Surveillance-Public-Use-Data/vbim-akqf.

4. He, X. et al. Temporal dynamics in viral shedding and transmissibility of COVID-19. Nature Medicine 26, 672–675 (2020).

5. Dong, E., Du, H. & Gardner, L. An interactive web-based dashboard to track COVID-19 in real time. The Lancet Infectious Diseases 20, 533–534 (2020).

6. Anderson, J. L. An Ensemble Adjustment Kalman Filter for Data Assimilation. Mon. Wea. Rev. 129, 2884–2903 (2001).

7. Shaman, J. & Karspeck, A. Forecasting seasonal outbreaks of influenza. Proceedings of the National Academy of Sciences 109, 20425–20430 (2012).

8. Yang, W., Lipsitch, M. & Shaman, J. Inference of seasonal and pandemic influenza transmission dynamics. PNAS 112, 2723–2728 (2015).

9. Pei, S., Kandula, S., Yang, W. & Shaman, J. Forecasting the spatial transmission of influenza in the United States. Proc Natl Acad Sci USA 115, 2752–2757 (2018).

10. Reis, J. & Shaman, J. Retrospective Parameter Estimation and Forecast of Respiratory Syncytial Virus in the United States. PLOS Computational Biology 12, e1005133 (2016).

11. DeFelice, N. B., Little, E., Campbell, S. R. & Shaman, J. Ensemble forecast of human West Nile virus cases and mosquito infection rates. Nature Communications 8, 14592 (2017).

12. Pei, S. & Shaman, J. Initial Simulation of SARS-CoV2 Spread and Intervention Effects in the Continental US. medRxiv 2020.03.21.20040303 (2020) doi:10.1101/2020.03.21.20040303.

13. Havers, F. P. et al. Seroprevalence of Antibodies to SARS-CoV-2 in 10 Sites in the United States, March 23-May 12, 2020. JAMA Intern Med (2020) doi:10.1001/jamainternmed.2020.4130.

14. CDC. Commercial Laboratory Seroprevalence Surveys. Centers for Disease Control and Prevention https://www.cdc.gov/coronavirus/2019-ncov/cases-updates/commercial-lab-surveys.html (2020).

15. Bajema, K. L. et al. Estimated SARS-CoV-2 Seroprevalence in the US as of September 2020. JAMA Intern Med (2020) doi:10.1001/jamainternmed.2020.7976.

16. CDC. Nationwide Commercial Laboratory Seroprevalence Survey. Centers for Disease Control and Prevention https://covid.cdc.gov/covid-data-tracker (2020).

17. Self, W. H. et al. Decline in SARS-CoV-2 antibodies after mild infection among frontline health care personnel in a multistate hospital network −12 states, April-August 2020. Morb. Mort. Weekly Rep. 69, 1762–1766 (2020).

18. Rogan, W. J. & Gladen, B. Estimating prevalence from the results of a screening test. American Journal of Epidemiology 107, 71–76 (1978).

19. Buss, L. F. et al. Three-quarters attack rate of SARS-CoV-2 in the Brazilian Amazon during a largely unmitigated epidemic. Science (2020) doi:10.1126/science.abe9728.

20. Yang, W. et al. Estimating the infection-fatality risk of SARS-CoV-2 in New York City during the spring 2020 pandemic wave: a model-based analysis. The Lancet Infectious Diseases (2020) doi:10.1016/S1473-3099(20)30769-6.

21. CDC. COVIDView, Key Updates for Week 1. Centers for Disease Control and Prevention https://www.cdc.gov/coronavirus/2019-ncov/covid-data/covidview/index.html (2021).

